# Resilliance Among Turkish Adolescents: A Multi-Level Approach

**DOI:** 10.1101/2024.02.25.24303348

**Authors:** Gökhan Çakir, Utku Işik, Umit Dogan Ustun, Nihan Su, Osman Gumusgul

## Abstract

**Introduction:** The objective of this study is to determine the components that contribute to psychological resilience in adolescents and to determine if physical exercise, emotion control, or self-efficacy are more effective predictors of resilience.

**Methods:** Data from participants was collected through a personal information form, the International Physical Activity Questionnaire—Short Form, the Self-Efficacy Scale for Children, the Emotion Regulation Scale for Children and Adolescents, and the Psychological Resilience Scale for Children and Adolescents. The data were gathered online from 16 out of the 81 provinces in Turkey, representing 7 different regions, using convenience sampling. The study sample comprised 505 adolescents, with 309 females and 196 males. The average age of the participants was 15.66 years, with a standard deviation of 1.34. The data obtained from the students was analyzed using SPSS 27.0 statistical software. The Chi-Square test was employed to establish the correlation between the demographic features of adolescents and their levels of physical activity. The relationship between the independent variables and the dependent variable was determined using correlation analysis and hierarchical regression analysis.

**Results:** The results suggest that physical exercise, the ability to regulate emotions through reappraisal, and self-efficacy are significant indicators of adolescents’ resilience.

**Conclusions:** The research conclusions point out that self-efficacy has a greater impact on psychological resilience compared to physical activity and emotion regulation.

## 1 INTRODUCTION

The mental well-being of adults is significantly influenced by their mental condition during childhood and adolescence (Mesman et al., 2021). Adolescence is commonly known as the “danger zone,” according to the World Health Organization (WHO, 2019), as cognitive, emotional, and behavioral growth may be affected as a result of difficult emotional reactions such as separation from parents, identity formation, and individuation (Lund et al., 2018; Selçuk, 2023). The mental well-being of teenagers is intricately connected to their social growth (Zhu et al., 2023). Today’s perpetually changing and evolving social environment has placed adolescents under unprecedented levels of stress (Caqueo-Urzar et al., 2023). The presence of feelings of rejection and judgment in social situations can diminish an individual’s confidence, potentially resulting in the development of more enduring issues in the future. During adolescence, individuals are susceptible to a range of mental and physical developmental vulnerabilities and dysfunctions, including mood disorders, substance misuse, and obesity (Hugh-Jones et al., 2022; Romeo, 2018). Hence, it is crucial to ascertain strategies for effectively managing the diverse problems and pathologies encountered by teenagers, with the aim of fostering a healthier transition into adulthood (Guo & Liang, 2023). Psychological resilience is a highly effective strategy for dealing with the difficulties of adolescence. Psychological resilience is positively and significantly associated with levels of mental health (Fastame et al., 2022; Chan et al., 2021). The study focuses on the dependent variable of psychological resilience, which is defined as the capacity to effectively adjust to and cope with life’s challenges and adversities (Las Hayas et al., 2019). Resilient individuals have the ability to ascribe favorable interpretations to intricate occurrences, manage adverse emotions, and adjust to evolving external stressors over the course of their lives (Xu et al., 2021). Insufficient levels of psychological resilience in teenagers can cause a delay in their mental and social development, as well as a lack of independence and responsibility. This can then contribute to the development of gaming addiction, excessive anxiety, academic pressure, and other related symptoms (Zhai and Ji, 2022). Additionally, it can result in melancholy, an incapacity to manage stress, suicide, subpar academic achievement, and other troublesome behaviors (Kalisch et al., 2017). Recent studies have observed a concerning rise in depression and anxiety among teenagers and young adults, leading to decreased levels of life satisfaction. These issues have notably intensified following the onset of the pandemic (Lattke et al., 2022; Preetz et al., 2021). At this point, it is crucial to develop resilience, as it is a determinant of the life satisfaction of adolescents. However, can resilience be developed? Selçuk (2023) argues that the psychological resilience of individuals is a dynamic construct, therefore necessitating an understanding of resilience as an evolving process. In other words, there are concepts that feed and develop the resilience of individuals in a wide range from childhood to adulthood, such as art, play, physical activity, self-efficacy, mindfulness, meditation, self-regulation, and emotion regulation. So, the present study aims to determine which of physical activity, emotion control, and self-efficacy is a more precise indicator of psychological resilience when these concepts are seen as determinants of individual resilience. Providing an explanation of the conceptual framework is beneficial in understanding the rationale behind the concentration on these three factors.

### 1.1 Theoretical background

#### 1.1.1 Physical activity and resilience

Physical activity has a positive impact on the social adjustment development of adolescents (Tan & Li, 2022). In addition, physical activity has been found to provide protection against mental health disorders such as depression and tension (Peyer et al., 2023; Szuhany et al., 2023). Evidence showing the beneficial impact of physical activity on mental health has consistently persisted in the literature throughout modern science. A study conducted with a sample of university students revealed that engaging in physical activity had a positive impact on psychological resilience (Li & Guo, 2023). While all forms of physical activity are recognized to be beneficial for overall well-being, individuals who participate in high-intensity exercise demonstrate greater psychological resilience, as observed by Szuhany et al. (2023). Current studies indicate that engagement in physical activity during adolescence might reduce unpleasant emotions. Engaging in regular physical activity can have a transformative effect on mental well-being, converting feelings of worry, despair, and stress into good emotions like enjoyment, joy, and relaxation (Malm et al., 2019; Nakajima et al., 2023). To clarify, encouraging physical activities that boost psychological resilience may serve as a means to increase the mental health of adolescents (Hu et al., 2015). Consistent with these studies, it is anticipated that the physical activity levels of adolescents may impact their resilience. Given the specifics, the subsequent hypothesis was formulated:

H1: The resilience of adolescents is significantly influenced by their levels of physical activity.

#### 1.1.2 Emotional regulation and self-efficacy as factors influencing psychological resilience

Emotional regulation is a crucial notion that promotes psychological resilience. Emotional regulation is a technique that aims to modify the influence of emotions on people (Tetik & Önder, 2021). The emotional regulation approach, as identified by Gratz and Roemer in 2004, is one of the frameworks that helps in managing emotions. Regulating one’s emotions in the face of challenging situations is seen as a crucial procedure for fostering optimal psychological functioning in adolescents (Park et al., 2020). The ability to regulate emotions might be seen as a valuable tool that protects adolescents from engaging in unreasonable and harmful behaviors (Mestre et al., 2017). Adolescents must prioritize the development and utilization of positive self-regulation skills in order to strengthen their psychological resilience (López-Valle, 2018). Reappraisal and suppression are two separate strategies highlighted in the literature on emotion regulation. Reappraisal is conceptualized as a cognitive change strategy, while suppression is considered a response modulation strategy. According to Tetik and Önder (2021), reappraisal strategies have been found to have adverse effects, leading to depressive symptoms. On the other hand, suppression strategies show a positive connection with these adverse effects. Consequently, re-appraisal strategies are thought to be linked to resilience. Self-efficacy is another concept that contributes to the development of psychological resilience (Li & Preziosi, 2022). Self-efficacy refers to an individual’s belief in their ability to perform well in a specific environment and achieve goals related to a task (VandenBos, 2007). Researchers have conceptualized self-efficacy as an expression of one’s general confidence in dealing with effort-demanding or novel situations (Telef & Karaca, 2012). At this juncture, a relationship between self-efficacy and resilience is postulated. The correlation between psychological resilience and self-efficacy in adolescence is widely regarded as an essential variable in promoting positive adaptation throughout this phase of growth. The link under examination assesses the impact of perceived self-efficacy on life skills, specifically in terms of overcoming problems (Sagone et al., 2020). Given the circumstances, the subsequent hypotheses have been formulated:

H2: The resilience of adolescents is impacted by emotion regulation strategies.

H3: The self-efficacy levels of adolescents have an impact on their resilience levels.

### 1.2 Present study

The primary objective of this study was to ascertain the extent to which self-efficacy, emotional regulation, and physical activity contribute to the development of psychological resilience in adolescents. Additionally, the study aimed to identify which variables better predict psychological resilience, as shown in Figure 1.

**FIGURE 1:**
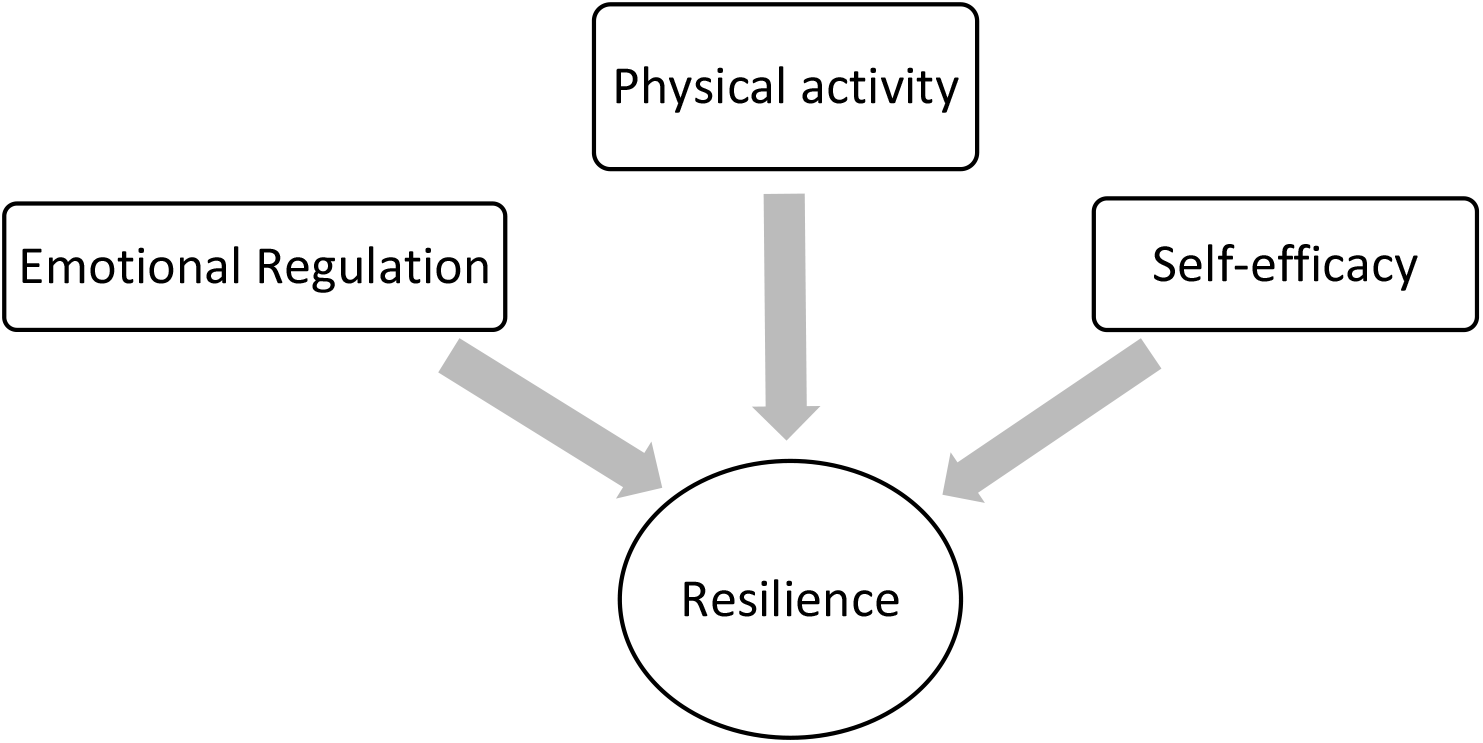
Research model.

Psychological interventions targeting adolescents’ resilience can reduce stress, improve their overall well-being and mental health, and cultivate their long-term coping abilities (Xing et al., 2023). Adolescence is a stage in life where young people are susceptible to mental and physical vulnerabilities and limitations in functioning. Thus, it is essential to cultivate elements that foster psychological resilience in order to maintain the overall well-being of adolescents, both physically and mentally. After conducting a thorough examination of the existing literature, only a few studies have been found that clarify and assess the correlation between physical activity and resilience by employing multiple variables. Expanding upon the concept that physical, mental, and emotional aspects may be interrelated, it is believed that addressing factors that can improve adolescents’ resilience in a comprehensive manner might address certain deficiencies in the existing literature. The objective of this study was to identify the factors that contribute to psychological resilience and ascertain whether physical activity, emotion control, or self-efficacy were better predictors of resilience.

## 2 MATERIAL AND METHOD

### 2.1 Research model

This research utilized the correlational survey model, commonly employed in quantitative research approaches. Correlational survey designs aim to determine the presence and/or degree of change between two or more variables (Büyüköztürk, 2015). The independent variables in this study were physical activity, self-efficacy, and emotion regulation. The dependent variable was resilience. Thus, the aim of this research model was to identify the determinants of psychological resilience and to determine which physical activity, emotion regulation, and self-efficacy serve as better predictors of resilience (Figure 1).

### 2.2 Participants, procedure, and ethics

Convenience sampling was employed in this study, which was cross-sectional in nature. G*Power (3.1.9.7, Faul et al., 2007) was used to determine the required sample size. G*Power is a free program that assists researchers in determining the practical sample size. According to G*Power, a sample of 439 participants is sufficient for three predictive variables with a real power of .95 (Deng et al., 2023; Nakajima et al., 2023). Additionally, for multivariate analyses (e.g., stepwise regression), a sample size 40 times the number of independent variables is deemed adequate (Tabachnick & Fidell, 2018).

During the research, data was collected from 621 participants. 18 individuals (2.8%) under the age of 14 and over 19, as well as 55 participants (9.1%) with incomplete or randomly filled questionnaires, were excluded from the analysis. Moreover, to identify outliers, the %5 trimmed value in the descriptive table was checked, resulting in the exclusion of 43 data points (7.8%) from the analysis. Consequently, data from 505 adolescent participants sampled conveniently were evaluated within the scope of the analysis.

Of the 505 participants, 196 (38.8%) were male, and 309 (61.2%) were female, with ages ranging from 14 to 19 years (15.65 ± 1.34). The number of participants engaged in sports and those not engaged in sports were 135 (26.7%) and 370 (73.3%), respectively. According to the International Physical Activity Questionnaire, 4.1% of the participants were found to have high physical activity levels. Table 1 displays the numerical distribution of participants based on their physical activity levels. Significant differences in physical activity levels based on sports engagement and gender were identified through chi-square test analyses (p<0.05). Participants with generally low and moderate physical activity levels were found to have lower participation in sports, while those with high physical activity levels had a higher rate of sports participation. Additionally, there were more male participants with low and moderate physical activity levels, whereas there were more female participants with high physical activity levels (p>0.05).

**TABLE 1.** Frequencies according to participants’ physical activity level.

|  |  | All<br>(%) | Low<br>(%) | Mid<br>(%) | High<br>(%) | p-value |
| --- | --- | --- | --- | --- | --- | --- |
| Class | Grade-1 | 25.545 | 23.232 | 26.744 | 25.000 | 0.287 |
|  | Grade-2 | 16.832 | 10.101 | 19.767 | 16.216 |  |
|  | Grade-3 | 18.416 | 20.202 | 16.279 | 20.946 |  |
|  | Grade-4 | 39.208 | 46.465 | 37.209 | 37.838 |  |
| Engaged in Sports | Yes | 26.733 | 11.111 | 14.729 | 58.108 | p<0.001 |
|  | No | 73.267 | 88.889 | 85.271 | 41.892 |  |
| Gender | Men | 61.188 | 84.848 | 65.116 | 38.514 | p<0.001 |
|  | Women | 38.812 | 15.152 | 34.884 | 61.486 |  |

### 2.3 Data collection tools

Personal Information Form: The researcher created this form to gather demographic data about the study participants’ gender, age, and level of physical activity.

Children’s Self-Efficacy Scale: This scale was developed by Muris (2001), and the Turkish adaptation study was conducted by Telef (2012). The Children’s Self-Efficacy Scale is a five-point Likert-type scale (1=not at all and 5=very well). Total self-efficacy sub-factor scores are calculated by summing the relevant items. The highest score that can be obtained from the scale is 105, and the lowest score is 21. A high score on the scale indicates a high level of self-efficacy related to children, and a low score on the scale indicates a low level of self-efficacy for children. When the internal consistency coefficients of the Children’s Self-Efficacy Scale were examined, a coefficient of .86 was calculated for the scale as a whole. In this study, the internal consistency coefficient was calculated as .87.

Child and Adolescent Emotion Regulation Scale: This scale was developed by Gullone & Taffe (2012), and the Turkish adaptation study was conducted by Tetik & Önder (2021). The sample of the scale consists of 1048 (558 girls and 490 boys) students who are studying in middle schools and high schools in the central districts of Adana province between 2017 and 2018. The scale consists of two separate dimensions, defined as reappraisal and suppression. The Cronbach alpha internal consistency coefficients of the sub-dimensions are .79 for the reappraisal sub-scale and .53 for the suppression sub-scale (Tetik & Önder, 2021). In this study, the internal consistency coefficient was calculated as .71 for the reappraisal sub-scale and .64 for the suppression sub-scale.

Child and Adolescent Psychological Resilience Scale (CAPS-12): Liebenberg et al. (2013) were responsible for developing this scale, and Arslan (2015) was in charge of the Turkish adaptation study. The study included a total of 256 students studying in middle school and high school in the centre of Isparta province. The ages of the participants range from 11 to 16. The mean age is 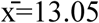 (SS=.97). The scale consists of 12 items and a single-factor structure. Within the scope of the reliability study, the internal consistency coefficient was calculated for the scale. The Cronbach alpha value was calculated as .91. In this study, the internal consistency coefficient was calculated as .80.

International Physical Activity Questionnaire (Short Form) (IPAQ): The questionnaire is a short form of the International Physical Activity Questionnaire. It was developed to determine the activity levels in the past 7 days. These days include the duration (in minutes) and number of days of sitting, walking, moderate-intensity activities, and vigorous activities. To calculate the points, multiply the MET (multiples of resting oxygen consumption) value by the day and minute. The MET value is calculated as 3.3 for walking, 4 for moderate-intensity activities, and 8 for vigorous activities (Savcı et al., 2006). Before classification, the scores for walking, moderate-intensity activities, and vigorous activities are determined, and the person’s total score is calculated. In the classification based on the calculated total score, the person is included in the inactive (low), minimal active (medium), and very active (high) groups if the <600 MET-minutes/week, 600-3000 MET-minutes/week, and >3000 MET-minutes/week, respectively (Savcı et al., 2006). In this study, only categorical MET values were used to determine the distribution of the participants, and all other analyses were conducted based on the total MET values.

### 2.4 Data Analysis

The data obtained from adolescents was analysed using the SPSS 27.0 package program. Descriptive statistics were used to reveal the demographic characteristics of the participants. In this stage, the mean and standard deviation values of all variables were calculated, and the normality assumption was checked by examining the skewness and kurtosis values. The ±2 base of George & Malley (2016) was taken as the reference value for the normality assumption.

The relationship between adolescents’ demographic characteristics and their physical activity levels was determined using a chi-square analysis. The impact of independent variables on the dependent variable was established through hierarchical regression analysis. Prior to conducting these analyses, assumptions were examined. For each model, regression analysis assumptions including normality, linearity, autocorrelation (Durbin-Watson test value = 2.70), constant variance (homoscedasticity), detection of outliers (DFBeta, Cook’s Distance), and multicollinearity were tested (Tabachnick & Fidell, 2018), and it was observed that the assumptions were met. Additionally, a correlation analysis was performed before the hierarchical regression analysis to determine the relationship between the variables.

## 3 RESULTS

### 3.1 Descriptive statistics and correlation analysis

Table 2 presents the means, standard deviations (SD), and Pearson’s correlation coefficients for Physical Activity, reappraisal, suppression, resilience, and self-efficacy. The results demonstrated a significant correlation between the study variables. Physical activity was significantly positively correlated with reappraisal (r = 0.214, p < 0.001), resilience (r = 0.220, p < 0.001), and self-efficacy (r = 0.251, p < 0.001). Reappraisal was significantly positively correlated with suppression (r = 0.089, p < 0.05), resilience (r = 0.411, p < 0.001), and self-efficacy (r = 0.463, p < 0.001). Resilience was significantly and positively correlated with self-efficacy (r = 0.582, p < 0.001). The significant correlations among the study variables provide initial support for the proposed hypotheses.

**TABLE 2.** Descriptive statistics and Pearson correlations of the study variables.

|  | 1 | 2 | 3 | 4 | 5 | 6 | 7 |
| --- | --- | --- | --- | --- | --- | --- | --- |
| 1. Gender | 1 |  |  |  |  |  |  |
| 2.Age | 0.057 | 1 |  |  |  |  |  |
| 3.Engaged in Sports | -.318** | -0.019 | 1 |  |  |  |  |
| 4.Physical Activity | .307** | 0.037 | -.461** | 1 |  |  |  |
| 5.Reappraisal | .125** | -0.014 | -.162** | .214** | 1 |  |  |
| 6.Resilience | .125** | -.095* | -.209** | .220** | .411** | 1 |  |
| 7.Self-efficacy | .247** | -0.024 | -.138** | .251** | .463** | .582** | 1 |
| Mean |  | 15.650 |  | 2681.09 | 20.74 | 45.51 | 66.38 |
| SD |  | 1.340 |  | 3194.11 | 3.54 | 6.84 | 13.25 |

Note: M = mean; SD = standard deviation. Gender is a dummy code (female = 1; male = 2). Engaged in Sports is a dummy code (yes= 1; no = 2). \* $p < 0.05$ , \*\* $p < 0.01$

**TABLE 3.** Analysis of hierarchical regression between variables.

| Dependent | Predictors | B | Std. Error | Beta | t | p |
| --- | --- | --- | --- | --- | --- | --- |
| Resilience<br>(Model 1) | (Constant) | 44.259 | 0.388 |  | 114.068 | 0.000 |
|  | Physical Activity | 0.000 | 0.000 | 0.220 | 5.049 | 0.000 |
| *R*=.220; *R*<sup>2</sup>=.048; *df*=1.503; *F*=25.495; *p*<0.01 |  |  |  |  |  |  |
| Resilience<br>(Model 2) | (Constant) | 29.490 | 1.637 |  | 18.018 | 0.000 |
|  | Physical Activity | 0.000 | 0.000 | 0.138 | 3.350 | 0.001 |
|  | Reappraisal | 0.734 | 0.079 | 0.381 | 9.249 | 0.000 |
| *R*=.432; *R*<sup>2</sup>=.187; *df*=1.502; *F*=85.539; *p*<0.01 |  |  |  |  |  |  |
| Resilience<br>(Model 3) | (Constant) | 21.606 | 1.587 |  | 13.613 | 0.000 |
|  | Physical Activity | 0.000 | 0.000 | 0.060 | 1.630 | 0.104 |
|  | Reappraisal | 0.331 | 0.078 | 0.172 | 4.260 | 0.000 |
|  | Self-efficacy | 0.252 | 0.021 | 0.488 | 11.981 | 0.000 |
| *R*=.607; *R*<sup>2</sup>=.368; *df*=1.501; *F*=143.541; *p*<0.01 |  |  |  |  |  |  |

The hierarchical regression analysis used to find the variables that could predict the resilience of the participants showed that physical activity fit into the first model, reappraisal from the emotion regulation sub-dimensions fit into the second model, and self-efficacy fit into the third model.

Based on the analysis presented in Table 2, it was found that in all three models, participants significantly predicted resilience values. Upon examining Model 1, it was observed that physical activity alone accounts for 4.8% of the total variance of resilience (R = 0.220; R2 = 0.048; df = 1,503; F = 25.495; p<0.01) and significantly predicts resilience. In Model 2, when reappraisal, a sub-dimension of emotion regulation, was included in the model, physical activity still predicted resilience to a certain extent (β = 0.142, p<0.05), but the main strength of the model was provided by reappraisal (β = 0.381, p<0.01). In other words, when reappraisal was included in the model, the impact level of physical activity on resilience decreased but still remained significant (R = 0.432; R2 = 0.187; df = 1,502; F = 85.539; p<0.01). In Model 3, self-efficacy was finally included in the model, and it was understood that this model accounted for 36.8% of the total variance in resilience (R = 0.611; R2 = 0.373; df = 1,503; F = 145.585; p<0.01) and significantly predicted resilience. The impact level of self-efficacy emerged to be higher than the impact level of the other variables (β = 0.488, p<0.01). Additionally, when self-efficacy was included in the model, the impact of physical activity on resilience disappeared (β = 0.060, p > 0.05).

To better understand the relationships between the variables that create significant differences in psychological resilience within the models and their interactions, Jeremy Dawson’s slopes were used. As evident from the graphs below, the most influential factor in resilience is self-efficacy.

**GRAPH 1.**
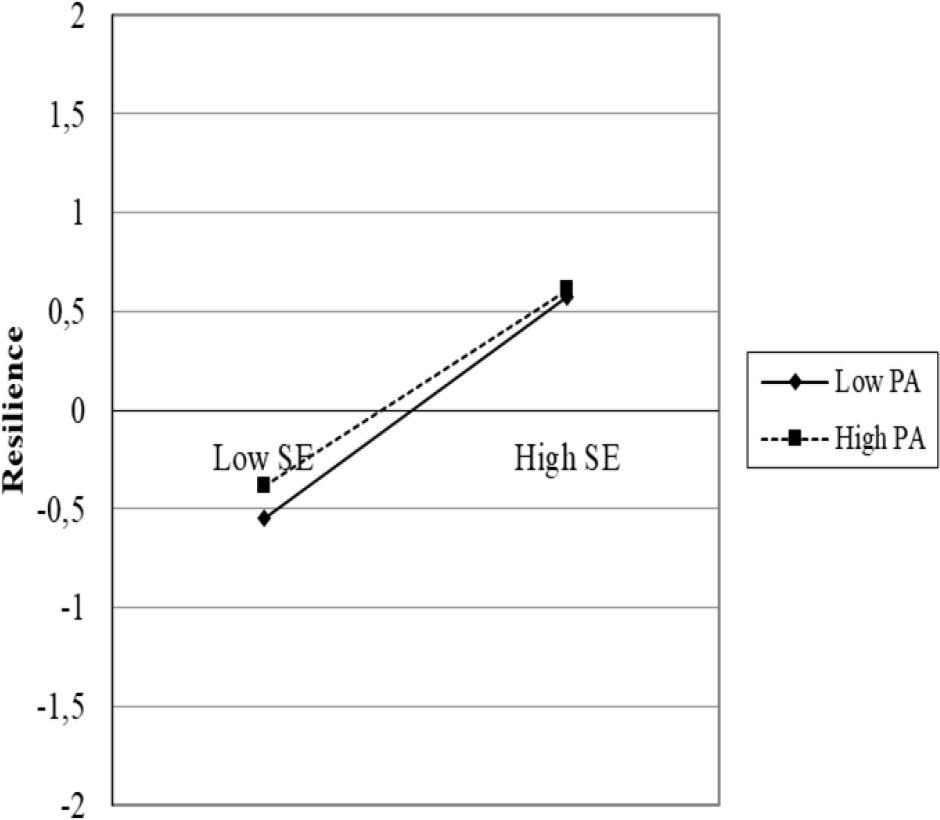
Self efficacy, physical activity and resilience.

**GRAPH 2.**
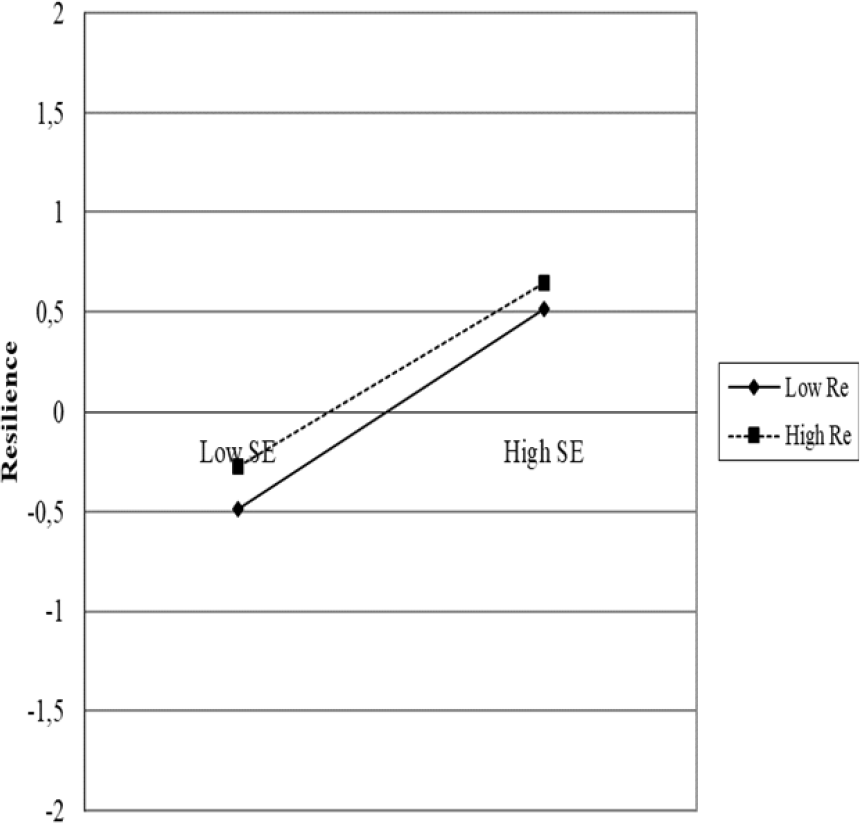
Self efficacy, reappraisal and resilience.

**GRAPH 3.**
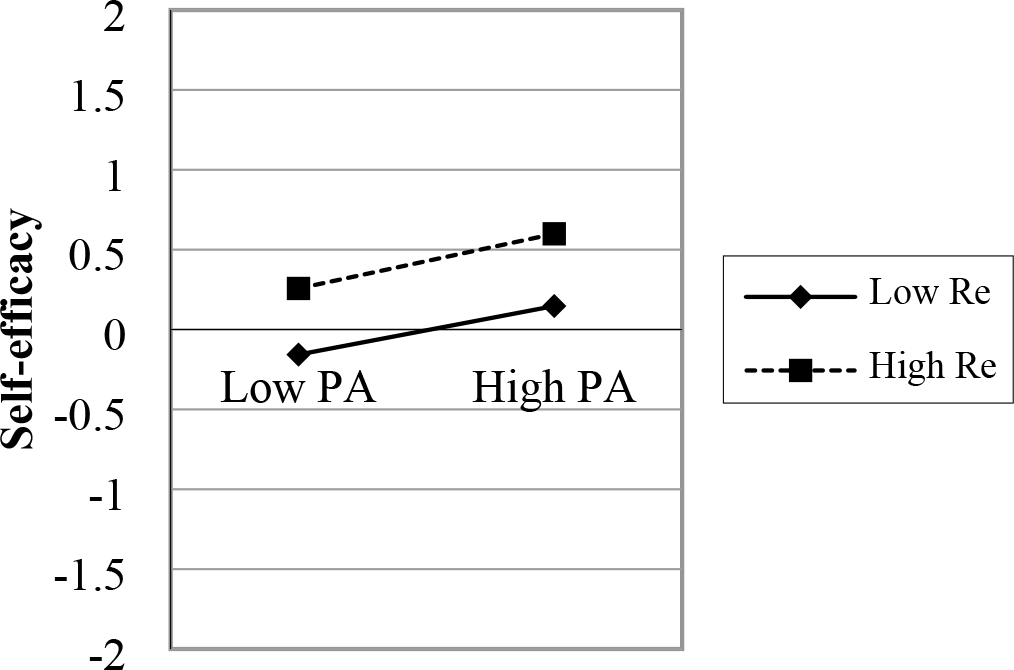
Self efficacy, physical activity and reappraisal. Not: PA= Physical Activity; SE: Self-efficacy; Re: Reappraisal

In the slopes showing the relationships between self-efficacy, physical activity, resilience, and reappraisal, it is evident that self-efficacy creates a noticeable difference between low and high levels. For instance, as observed from Graph 1 and Graph 2, a significant difference was noticed between low and high scores of self-efficacy (p<0.05), while low and high levels of physical activity and reappraisal showed similar changes. In other words, individuals with both low and high levels of physical activity and reappraisal who also exhibited high self-efficacy experienced increased resilience. The levels of physical activity and reappraisal did not create a significant difference in resilience (p<0.05). Indeed, Graph 3 supports this observation. It appears that physical activity and reappraisal are not as influential on resilience as self-efficacy.

## 4 DISCUSSION

During adolescence, individuals are prone to various mental and physical developmental vulnerabilities, which can be seen as constraints to maintaining a healthy lifestyle. One of the factors that promotes the physical and mental health of adolescents is psychological resilience. Through resilience, individuals can cope with negative emotions and adapt to changing external stressors throughout their lives. Therefore, interventions aimed at psychological resilience for adolescents can alleviate stress, enhance their well-being, and improve their mental health (Xing et al., 2023). By addressing mechanisms that enhance resilience and examining the relationships between these mechanisms, the development of a healthier generation can be promoted in every aspect. This research was conducted to identify mechanisms that enhance resilience in adolescents and to determine which variables better predict resilience.

The results of the study revealed that physical activity, emotion regulation (reappraisal), and self-efficacy significantly predicted resilience. This finding aligns with the idea that promoting physical activities that enhance resilience can improve the mental health of adolescents (Ho et al., 2015). In other words, it can be said that by participating in activities that can be performed at low, moderate, and high levels (such as folk dances, dancing, bowling, tennis, basketball, walking, and cycling), adolescents can better focus on completing tasks and maintain healthier relationships with friends, family, and at school.

It is crucial for adolescents to develop and utilise positive self-regulation strategies to enhance their resilience (López-Valle, 2018). Indeed, according to the research results, adolescents’ reappraisal regulation significantly predicted their resilience. This result suggests that adolescents who can think differently when feeling bad or happy may be more psychologically resilient.

Another finding indicates that adolescents’ self-efficacy influences their resilience. This finding can be interpreted as indicating that adolescents with higher beliefs in their academic, social, and emotional capacities are more resilient. This finding is consistent with Sagone et al. (2020), who emphasised the significant role of self-efficacy in life skills such as coping with challenges.

This study once again demonstrates the effects of physical activity participation, emotion regulation strategies, and self-efficacy on resilience, in line with the literature. What distinguishes this study from the existing literature is the varying levels of impact of the variables on resilience and how the inclusion of variables in sequence changes the effectiveness of one another. Both the analyses and the drawn slopes demonstrate that self-efficacy is more effective for psychological resilience than physical activity and emotion regulation. Although physical activity and emotion regulation strategies also have an impact on resilience, this impact is not as strong as self-efficacy. Additionally, when self-efficacy is included in the model (Model 3), the impact of physical activity on resilience disappears, indicating that physical activity alone cannot be a predictor without self-efficacy. Of course, we cannot disregard the impact of physical activity on resilience; this has already been demonstrated in both the existing literature and the findings of this study. However, without self-efficacy, no change can be made in psychological resilience, no matter how much physical activity is included. Graph 1 shows that low and high levels of physical activity show similar changes in psychological resilience, but there is a noticeable difference between low and high self-efficacy. The same applies to emotion regulation. Although the impact of emotion regulation on resilience continues in Model 3, as understood from Graph 2, the presence of low or high reappraisal does not cause any change in psychological resilience compared to low or high self-efficacy. Self-efficacy, emotion regulation, physical activity participation, and psychological resilience are all linked in a detailed and organised way in studies that look at these relationships. When these studies are looked at in this way, it’s easier to understand what we’ve found (Kim et al., 2023; Deng et al., 2023; Wu et al., 2022; Neumann et al., 2022). No matter how good an individual is at participating in physical activities or regulating their emotions, they cannot be psychologically resilient without developing good self-efficacy.

Finally, it should be noted that this study has some limitations. Although data were collected from 16 different provinces across the country, convenience sampling has its limitations. The cross-sectional nature of this study means that the data was obtained within a specific time frame. Therefore, there is a need for more comprehensive and longitudinal studies examining the determinants of psychological resilience. Finally, despite excluding incomplete and erroneous data from the analysis, the subjective nature of the physical activity questionnaire may lead to bias. This limitation can be considered an important constraint regarding the generalizability of the data.

## Data Availability

All data files are available from the Google Drive database. https://drive.google.com/drive/u/0/home

## Acknowledgements

We would like to thank the all adolescents who filled out the data collection tools completely so that this study could be carried out.

## CONFLICT OF INTEREST STATEMENT

The authors declare no conflict of interest.

## DATA AVAILABILITY STATEMENT

The data that support the findings of this study will be openly available upon acceptance of the manuscript in the Open Science Framework (OSF).

## ETHICS STATEMENT

This study was performed in line with the principles of the Declaration of Helsinki. Approval was granted by the University of …………………. University Social Sciences Ethic Board.

## Notes

### Competing Interest Statement

The authors have declared no competing interest.

### Funding Statement

The author(s) received no specific funding for this work.

### Author Declarations

This study was performed in line with the principles of the Declaration of Helsinki. Approval was granted by the University of Recep Tayyip Erdogan University Social Sciences Ethic Board. Approval Number and Date: 2023/45 / 27.09.2023

